# Efficacy and safety of *Anas barbariae* 200K in influenza-like illness: a randomized double-blind, placebo-controlled, multicenter study

**DOI:** 10.64898/2026.09.01.26361819

**Authors:** Manoj Gupta, Ashish Tajne, Pravin Chavan, Isabelle Chanel, Stéphanie Chanut, Naoual Boujedaini, Monica Gupta

## Abstract

**Objective:** Influenza-like illness (ILI) is characterized by the sudden onset of systemic and respiratory symptoms such as fever, fatigue and myalgia, requiring prompt management. *Anas barbariae* 200K, a homeopathic medicinal product, is traditionally used to relieve flu-like symptoms. This study aimed to evaluate its efficacy in alleviating ILI symptoms within 72 hours. Secondary outcomes assessed symptoms relief, adverse effects, functional outcomes, compliance, and participant and investigator satisfaction.

**Methods:** This prospective, randomized, double-blind, placebo-controlled multicenter superiority trial with two parallel groups was conducted in primary care in India in 2021. Children (6–12 years) and adults (18–65 years) with ILI symptoms for less than 24 hours were randomized to receive either *Anas barbariae* 200K (Oscillococcinum®) or placebo. Treatment was given three times daily for three days. The primary endpoint was the proportion of participants with alleviation of ILI symptoms within 72 hours following the initiation of treatment.

**Results:** A total of 707 participants received *Anas barbariae* 200K (n = 352) or placebo (n = 355). Within 72 hours, the *Anas barbaria*e 200K group showed significantly greater symptom relief than the placebo group (74% *vs.* 44.5%; *p* < 0.0001). Secondary endpoints also favored *Anas barbariae* 200K group, including faster symptom alleviation (46.7 *vs.* 67.3 hours; *p* < 0.0001) and shorter median time, estimated by KM analysis, to return to daily activities (4.0 *vs*. 6.0 days; *p* < 0.0001). No serious adverse events were reported.

**Conclusion:** *Anas barbariae* 200K significantly alleviated ILI symptoms within 72 hours, with improvements that were both statistically and clinically significant. Significant benefits were also observed across secondary outcomes, with no safety signal identified. This study showed that *Anas barbariae* 200K could be an effective and well-tolerated treatment for ILI symptoms.

**Trial registration:** Clinical Trials Registry of India, [CTRI](https://ctri.nic.in/Clinicaltrials/pmaindet2.php?EncHid=NTE2NDU=&Enc=&userName=): 2021/02/031250, registered on February 12, 2021.

## 1. Introduction

Influenza-like illness (ILI) refers to a clinical syndrome characterized by a sudden onset of systemic symptoms accompanied by respiratory symptoms.^1^ Systemic symptoms peak within 24 hours and diminish over the course of 3–7 days, while respiratory symptoms typically develop 24 hours after the onset of systemic symptoms and follow a similar pattern.^2^ Influenza-like syndrome is most frequently associated with respiratory viruses or subsequent bacterial infections.^3^ Because ILI symptoms are not pathogen-specific and cannot be reliably attributed to a particular infectious agent without laboratory confirmation, available epidemiological data largely focus on influenza viruses, which remain the most routinely tested pathogens.^4,5^

Annual fluctuations in ILI incidence challenge public health preparedness and may increase the risk of adverse outcomes in vulnerable populations. The morbidity, mortality, and economic burden of ILI are influenced by factors such as age, concomitant diseases, vaccination status, and the incidence and severity of secondary complications, and have been the focus of numerous studies.^6–10^ Elderly, children, and immunocompromised individuals are at greatest risk of complications. While antiviral therapy may be indicated for those patients, most uncomplicated cases are managed with symptomatic care aimed at relieving discomfort and supporting recovery. Regardless of patient profile, prevention and early therapeutic management can reduce symptom severity, facilitate faster recovery, preserve patients’ quality of life, and reduce the overall burden on health care systems.^11–13^

Additionally, many patients seek traditional, complementary, and integrative medicines (TCIM) approaches, such as homeopathic medicine.^14^ Previous observational studies conducted in France indicated that the most often recommended therapies were antipyretics such as paracetamol, oral or nasal sprays, combined treatments (allopathic and homeopathic), and homeopathy.^15,16^

*Anas barbariae* 200K is a homoeopathic medicinal product containing a single active ingredient prepared from a specific extract of the heart and liver from *Anas barbariae* (duck breed).^17^ *Anas barbariae* 200K has been registered by competent authorities based on its traditional use for relief of flu-like symptoms such as fever, headache, chills, and body aches at the onset of ILI symptoms or shortly after the full clinical presentation has emerged.^18–20^ Today, *Anas barbariae* 200K is marketed as an over-the-counter product or after prescription by a physician.^21^

The clinical efficacy of *Anas barbariae* 200K for the treatment of ILI has been evaluated in several clinical studies. Two placebo-controlled trials reported faster symptom resolution at 48 hours with *Anas barbariae* 200K (Ferley 1989: *p* = 0.03; Papp 1998: *p* = 0.023), though a subsequent Cochrane review identified methodological limitations and deemed evidence insufficient for definitive clinical recommendations. Safety analyses from the Ferley and Papp clinical studies did not identify any significant tolerability concerns, with *Anas barbariae* 200K demonstrating a safety profile comparable to placebo. Within this context and given the global interest in TCIM and patient-centered care, the present study aims to further evaluate data on the efficacy and safety of *Anas barbariae* 200K in treating ILI, compared with placebo, to isolate the specific therapeutic effect of the treatment.^26^

The primary aim was to evaluate the efficacy of *Anas barbariae* 200K in alleviating ILI symptoms within 72 hours of first intake in primary care settings in children and adults with ILI symptoms for less than 24 hours. The secondary objectives were assessing symptom relief, functional outcomes, tolerability and safety, compliance, participant and investigator satisfaction.

The protocol was reviewed and approved by the Royal Pune Independent Ethics Committee, the Independent Ethics Committee Healthpoint Hospital, the Institutional Ethics Committee Panchsheel Hospital, the Institutional Ethics Committee Narayana Diagnostic, and the Vagus Institutional Ethics Committee for this work.

## 2. Materials and Methods

### 2.1. Study design

This clinical trial was a randomized, double-blind, placebo-controlled, parallel-group, multicenter superiority study. The trial was implemented through general practitioners at ten independent primary care centers across India. All participants provided written informed consent prior to enrolment. The study included a three-day treatment phase and a seven-day follow-up period, with three scheduled visits: Day 1 (Visit 1), Day 3 (Visit 2), and Day 10 (Visit 3). At Visit 1, participants were screened for eligibility, randomized to receive either active treatment ( *Anas barbariae* 200K) or placebo and received the first dose of study medication at the clinic. The study was registered with the Clinical Trials Registry – India (CTRI/2021/02/031250) and reported in accordance with the CONSORT 2025 statement.

Participants recorded symptoms, treatment intake, concomitant medications, and daily activities in a diary throughout the study (three times daily from Day 1 to Day 3 then twice daily). Paracetamol was permitted as rescue medication, whereas other antipyretics, decongestants, antibiotics, antivirals, herbal remedies for influenza and immunosuppressive agents were prohibited and considered protocol deviations. At each visit, investigators reviewed symptoms, concomitant medications and safety, including serious adverse events (SAEs). At the final visit, both investigators and participants assessed treatment satisfaction.

### 2.2. Study population

Individual participants were eligible for inclusion if they were aged 6–12 years (pediatric) or 18–65 years (adult), presented with sudden onset of ILI symptoms within 24 hours, and were able to take the first dose of study medication within this time frame. ILI was defined as the presence of at least one systemic symptom (fever ≥37.8°C, malaise, headache, or myalgia) and one respiratory symptom (cough, sore throat, or shortness of breath). Participants were enrolled at the point of medical consultation for ILI symptoms, when disease severity was assessed and documented by the investigator. At baseline, investigators classified symptom severity as mild, moderate, or severe based on the participant’s most pronounced reported symptom, without the use of a standardized severity scale. Symptom alleviation was defined as an improvement of at least one severity level for every symptom documented at baseline. Inclusion and exclusion criteria are provided in **Supplemental Material**.

### 2.3. Randomization

Participants were randomly assigned 1:1 to receive either the active treatment or placebo using a centralized Interactive Web Response System (IWRS) PageOne™ by the investigator. The randomization schedule was generated by an independent statistician using computer-generated permuted blocks and uploaded to the IWRS, which linked treatment numbers to study medication. Randomization was performed based on pre-specified stratification by age group and the severity of ILI symptoms at baseline according to the most severe symptom since ILI started. Treatment allocation was concealed through the IWRS until randomization.

### 2.4. Intervention

Allocated treatments, *Anas barbariae* 200K (Oscillococcinum®), or placebo, provided by Laboratoires Boiron (France), were dispensed to participants at the initial visit. The treatment regimen consisted of three doses per day (morning, noon, and evening), for a total of nine doses, as scheduled, even if participants were symptom-free. Each dose was administered sublingually. Both participants and investigators were blinded to treatment allocation. The active treatment and placebo were visually indistinguishable in terms of appearance, taste, and packaging.

### 2.5. Primary and secondary outcomes

The primary outcome was the proportion of participants achieving ILI symptom alleviation at 72 hours after treatment initiation. Secondary outcomes are listed in **Table 1**. Safety outcomes included the occurrence of adverse events (AEs) and SAEs, collected from randomization until Visit 3 through spontaneous reporting by participants and monitoring by investigators. Events were coded using the Medical Dictionary for Regulatory Activities (MedDRA), version 23.1.

**Table 1.**
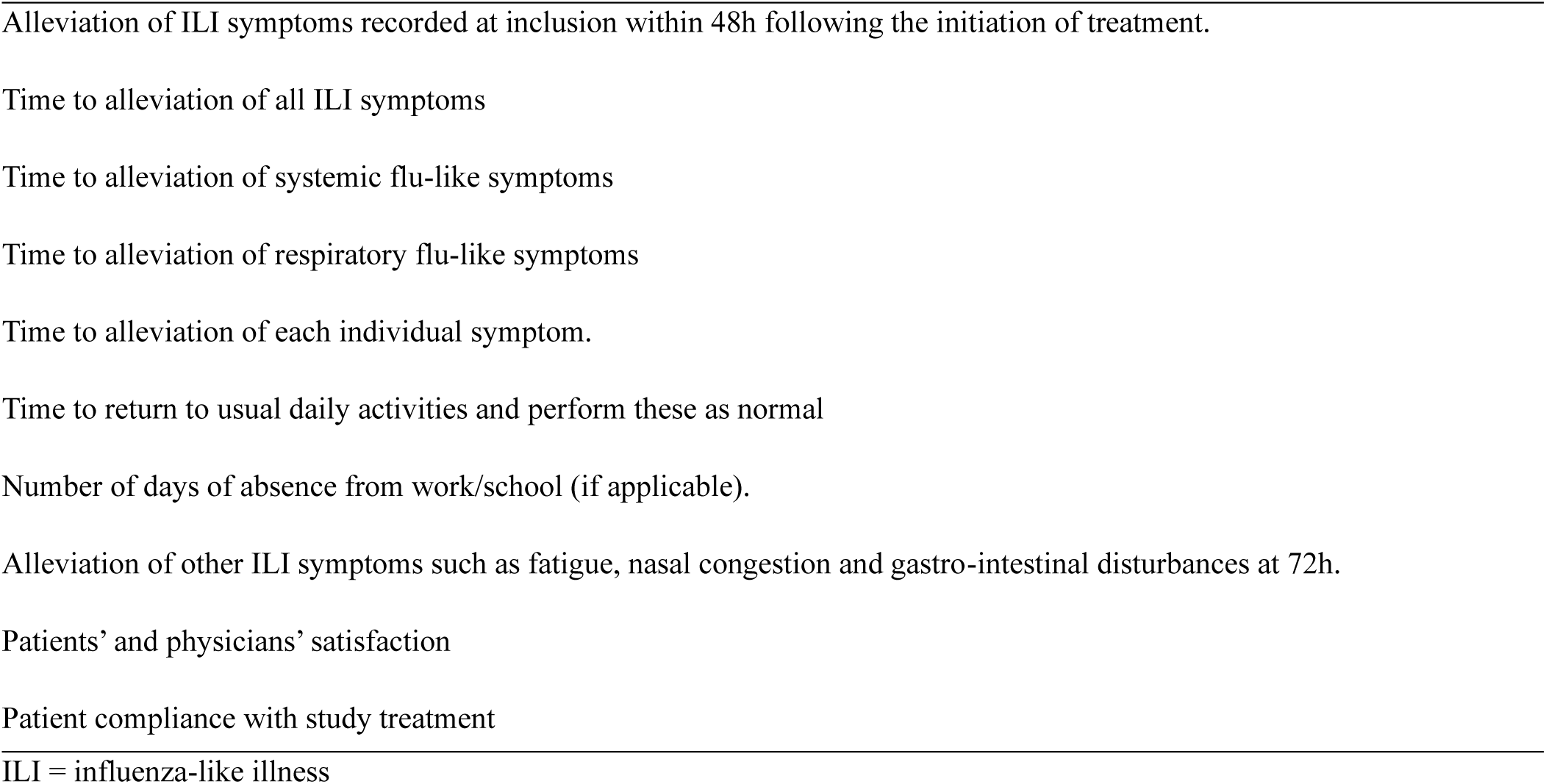
Secondary outcomes.

### 2.5. Statistical analysis

The sample size was calculated assuming that 80% of placebo-treated participants would achieve symptom alleviation at 72 hours. A total of 694 participants (347 per group) provided 80% power to detect a clinically meaningful absolute difference of 7% between groups using a one-sided alpha level of 0.05.

All efficacy results reported herein are based on the intention-to-treat (ITT) population; per-protocol (PP) analyses yielded consistent conclusions. The safety population comprised all participants who received at least one dose of study treatment. All statistical analyses were conducted using SAS software version 9.4. Results included parameter estimates, p-values, odds ratios (OR), and confidence intervals (CIs). Additional statistical details, including analysis populations, protocol deviations and statistical methods, are provided in the **Supplemental Material**.

## 3. Results

The study was carried out between March and September 2021 and enrolled both pediatric and adult populations. Both ITT and safety populations comprised 707 participants. Fifty participants (8 with critical and 42 with major protocol deviations) were excluded from the PP population, which comprised 657 individuals (92.9%) (**Fig. 1**). Definitions of critical and major deviations are provided in the **Supplemental Material**.

**Figure 1.**
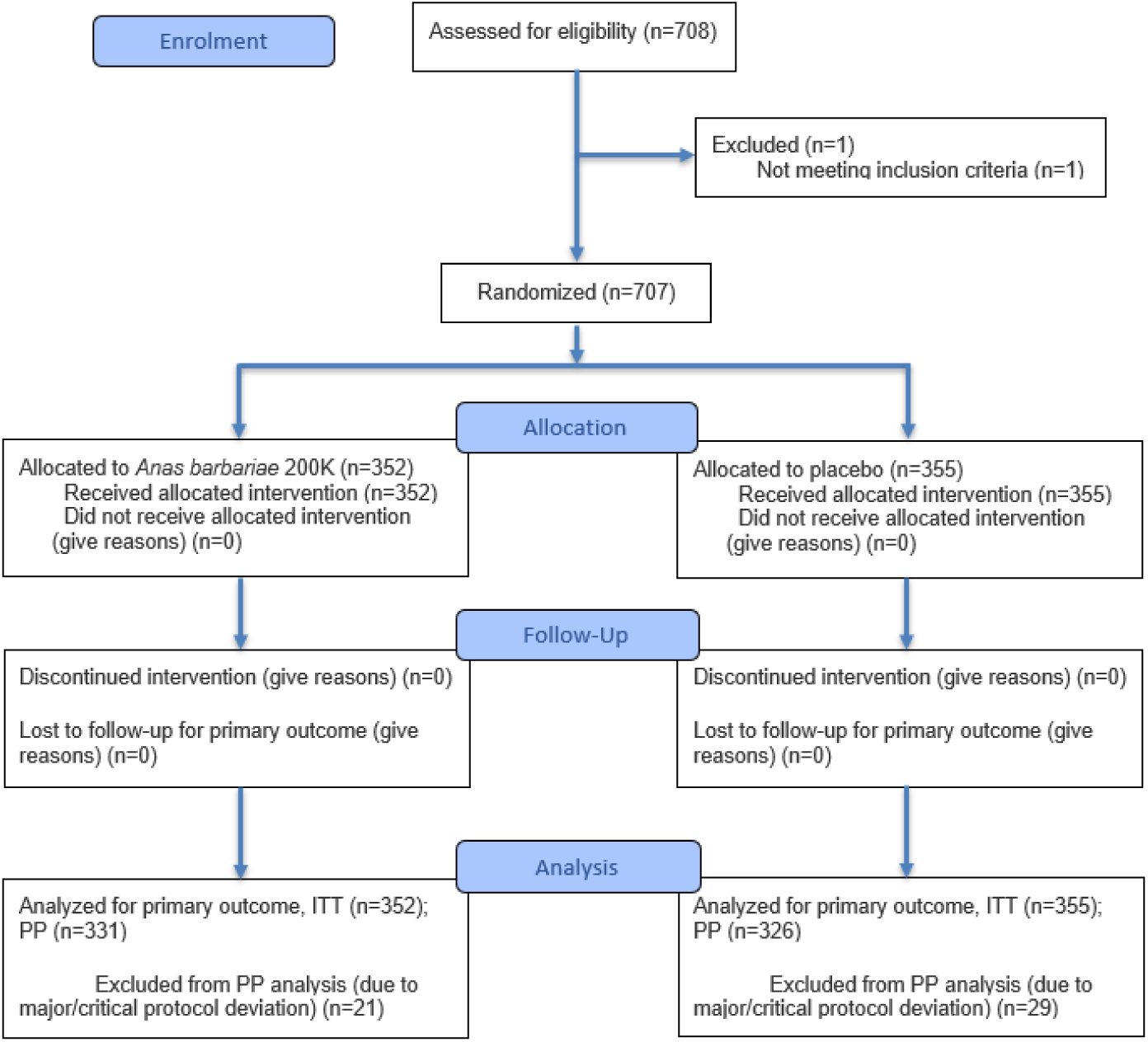
CONSORT Flow Diagram. ITT = intention-to-treat; PP = per protocol.

### 3.1. Population demographics

Demographic and clinical characteristics were comparable between groups at baseline (**Table 2**). Most participants presented with moderate or severe ILI symptoms. Influenza and pneumococcal vaccination rates were low and balanced between groups.

**Table 2.** Baseline characteristics of the study population by treatment group (ITT population)

| Characteristics |  | *Anas barbariae*<br>200K<br>(N = 352) | Placebo<br>(N = 355) |
| --- | --- | --- | --- |
| **Age (years)** |  |  |  |
|  | Mean (SD) | 26.2 (15.60) | 26.8 (16.45) |
|  | Median (min, max) | 26.0 (6, 63) | 27.0 (6, 65) |
| **Age group, n (%)** |  |  |  |
|  | Pediatric group (6-12years) | 121 (34.4%) | 123 (34.6%) |
|  | Adult group (18-65 years) | 231 (65.6%) | 232 (65.4%) |
| **Severity at baseline, n (%)** |  |  |  |
|  | Mild | 38 (10.8%) | 54 (15.2%) |
|  | Moderate | 195 (55.4%) | 164 (46.2%) |
|  | Severe | 119 (33.8%) | 137 (38.6%) |
| **Sex, n (%)** |  |  |  |
|  | Female | 142 (40.3%) | 150 (42.3%) |
|  | Male | 210 (59.7%) | 205 (57.7%) |
| **BMI, (kg/m<sup>2</sup>)** |  |  |  |
|  | Mean (SD) | 22.07 (4.182) | 22.34 (4.351) |
|  | Median (min, max) | 22.50 (10.5, 37.1) | 22.31 (10.2, 47.3) |
| **Vaccination, n (%)** |  |  |  |
|  | Influenza (2020-2021 season) | 2 (0.6%) | 3 (0.8%) |
|  | *Pneumococcus* | 23 (6.5%) | 20 (5.6%) |

BMI = body mass index; ITT = intention-to-treat; N = number of participants in specified treatment group; n = number of participants in a specified category; min = minimum; max = maximum; SD = standard deviation.

### 3.2. Primary endpoint

Participants in the active treatment group demonstrated significantly greater alleviation of all ILI symptoms within 72 hours of treatment initiation compared with the placebo group (74.0% *vs.* 44.5%; *p* < 0.0001) **(Fig. 2A**, **Table 3)**. This finding supports the statistically and clinically significant superiority of the active treatment over placebo in achieving quicker symptom relief. Subgroup analyses by age and baseline severity were consistent with the primary finding **(Fig. 2B-C).**

**Figure 2.**
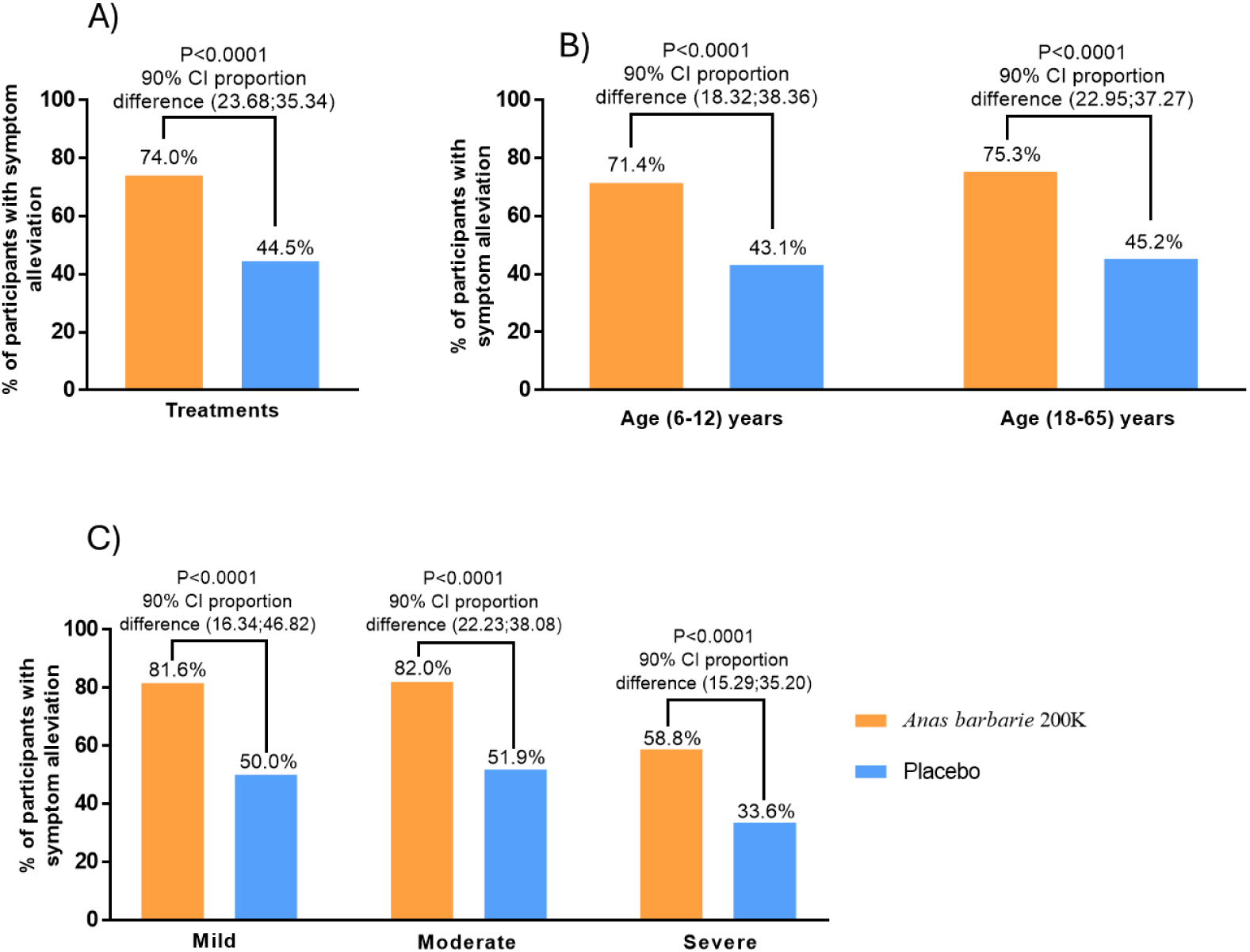
Percentage of participants (ITT analysis set) with alleviation of all ILI symptoms within 72 hours of treatment initiation (A) by treatments; (B) by age group; and (C) by severity of ILI symptoms. For primary endpoint (A), the percentages are based on the number of participants in the total population in the treatment groups, and a one-sided P-value was derived from the chi-squared test (superiority test). Error bars indicate the 90% CI for the point estimate. The 90% CI for the proportion difference was calculated using the normal approximation method without stratification. All analyses are based on the observed case analysis, with imputation for missing data. Stratification factors include the participants’ age groups and baseline severity. For subgroup analyses (B) and (C), the percentages are based on the number of participants in the subgroup population, and a one-sided P-value was derived from the chi-squared test. The 90% CI for the proportion difference was calculated using the normal approximation method without stratification. CI = confidence interval; ILI = influenza-like illness; ITT = intention-to-treat.

**Table 3.** Logistic regression analysis for alleviation of all ILI symptoms within 72 hours of intake of treatment (ITT population)

| **Statistic** | ***Anas barbariae* 200K**<br>**(N = 352)**<br>**n (%)** | **Placebo**<br>**(N = 355)**<br>**n (%)** | **ITT Population**<br>**(N=707)** |
| --- | --- | --- | --- |
| Subjects with alleviation of all ILI symptoms within 72 hours | 256 (74.0%) | 157 (44.5%) |  |
| Subjects with no alleviation within 72 hours | 90 (26.0%) | 196 (55.5%) |  |
| Missing | 6 | 2 |  |
| **Treatment Group (*Anas barbariae* 200K vs. Placebo)** |  |  |  |
| *P*-value |  |  | <.0001 |
| Odds ratio |  |  | 3.603 |
| 90% CI of the Odds Ratio |  |  | (2.732; 4.752) |
| **Age Group (Adult group vs. Pediatric group)** |  |  |  |
| *P*-value |  |  | 0.7320 |
| Odds ratio |  |  | 0.942 |
| 90% CI of the Odds Ratio |  |  | (0.707; 1.255) |
| **Symptoms Severity (Mild vs. Severe)** |  |  |  |
| *P*-value |  |  | 0.4724 |
| Odds ratio |  |  | 1.910 |
| 90% CI of the Odds Ratio |  |  | (1.207; 3.024) |
| **Symptoms Severity (Moderate vs. Severe)** |  |  |  |
| *P*-value |  |  | 0.0011 |
| Odds ratio |  |  | 2.510 |
| 90% CI of the Odds Ratio |  |  | (1.868; 3.372) |
| **Use of Paracetamol (No vs. Yes)** |  |  |  |
| *P*-value |  |  | 0.0193 |
| Odds ratio |  |  | 1.519 |
| 90% CI of the Odds Ratio |  |  | (1.132; 2.038) |

CI = confidence interval; ILI = influenza-like illness; ITT = intention-to-treat; N = number of participants in specified treatment group; n = number of participants in a specified category. The *p*-value, odds ratio and its 90% confidence interval (CI) will be presented comparing *Anas barbariae* 200K to placebo based on logistic regression model with all ILI symptoms alleviation as the outcome variable and treatment group as the factor and age group, symptoms severity, use of paracetamol as adjustments variables.

Multivariable regression analysis showed that participants in the active treatment group were 3.6 times more likely to experience symptom alleviation within 72 hours compared with the placebo group (OR = 3.603 [90% CI: 2.732, 4.752]; *p* < 0.0001), as shown in **Table 3**. Additionally, covariate analysis showed that age group (adult vs. pediatric) was not a significant predictor of symptom alleviation (OR = 0.942 [90% CI: 0.707; 1.255]; *p* = 0.7320). Analyses of symptom severity revealed no differences between participants with mild versus severe symptoms (OR = 1.910 [90% CI: 1.207, 3.024]; *p* = 0.4724). Participants presenting moderate symptom severity were 2.5 times more likely to achieve symptom alleviation than those with severe symptoms (OR = 2,510 [90 % CI: 1.868, 3.372]; *p* = 0.0011). Furthermore, paracetamol use was identified as a statistically significant covariate in the model; participants who reported using paracetamol were about 1.5 times more likely to achieve complete symptom alleviation within 72 hours compared with those who did not (OR = 1.519 [90% CI: 1.132, 2.038]; *p* = 0.0193) (**Table 3)**. Paracetamol use was shown to be similar between the two groups (54.8% (n=193) *vs*. 55.8% (n=198); *p* = 0.4002) (**Supplemental Material, Table S.1**).

### 3.3. Secondary endpoints

#### 3.3.1. Alleviation of ILI symptoms within 72 hours, maintained for 24 hours, and Alleviation of ILI symptoms recorded within 48 hours of treatment initiation

A higher percentage of participants in the active treatment group experienced relief from all ILI symptoms within 72 hours, lasting at least 24 hours, compared with the placebo group (72.8% *vs.* 41.1%; *p* < 0.0001). Additionally, the active treatment was associated with faster symptom relief within 48 hours compared with placebo (42.2% *vs.* 25.4%; *p* < 0.0001) (**Table 4**).

**Table 4.**
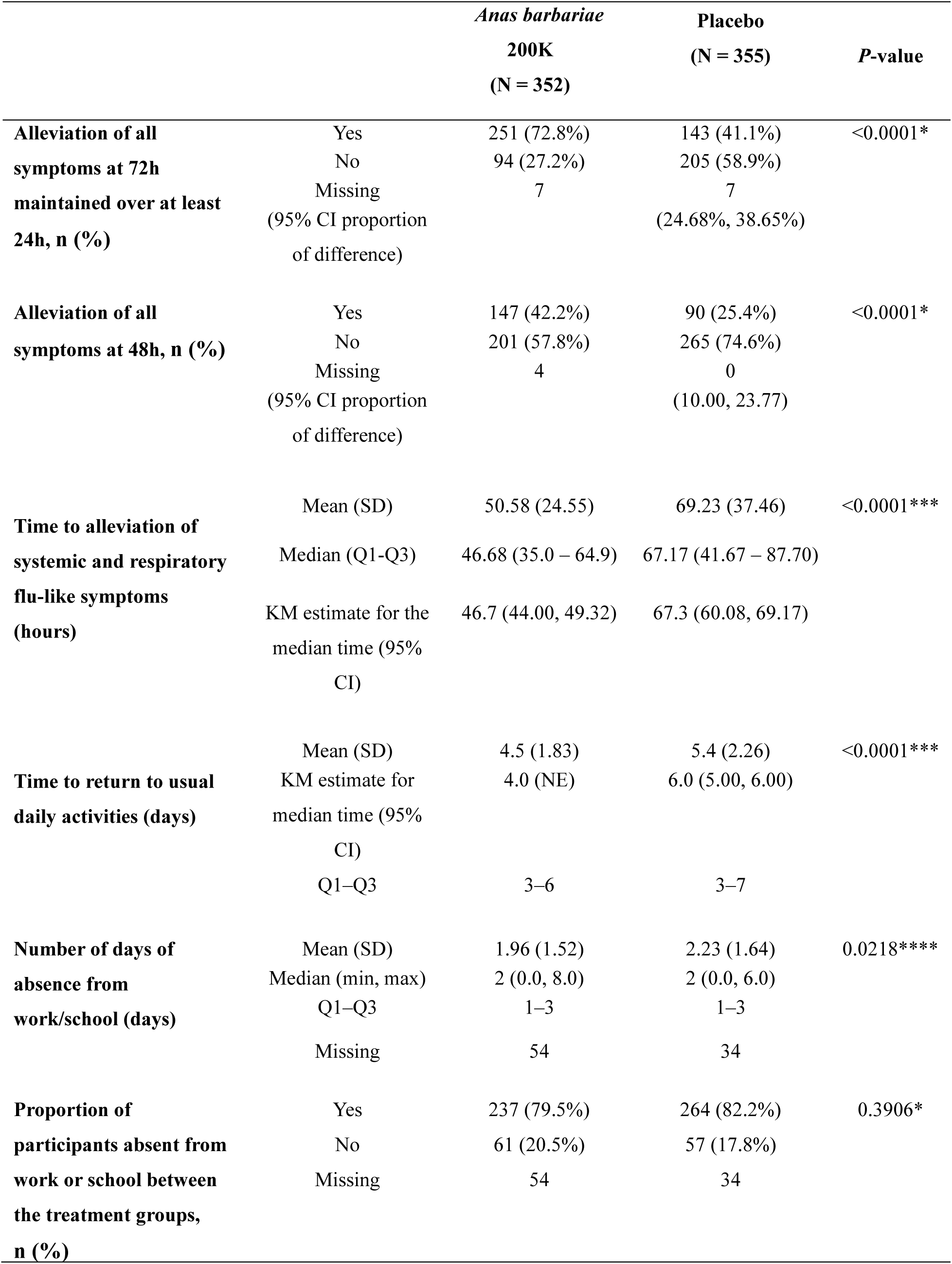
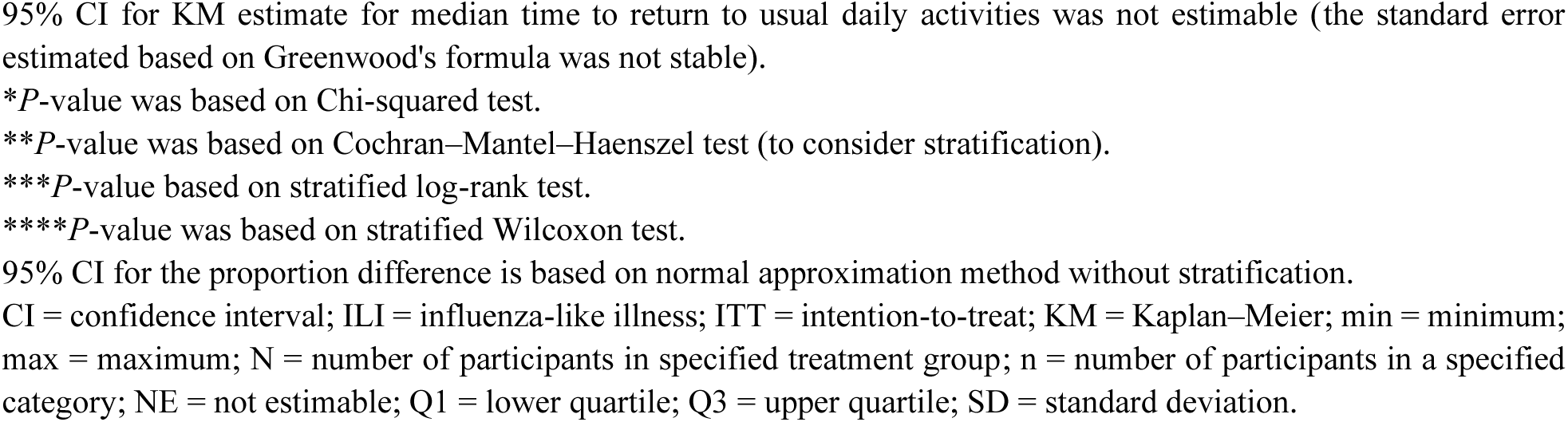
Summary of secondary endpoints related to alleviation of ILI symptoms (ITT population)

|  |  | *Anas barbariae*<br>200K<br>(N = 352) | Placebo<br>(N = 355) | P-value |
| --- | --- | --- | --- | --- |
| **Alleviation of all symptoms at 72h maintained over at least 24h, n (%)** | Yes | 251 (72.8%) | 143 (41.1%) | <0.0001* |
|  | No | 94 (27.2%) | 205 (58.9%) |  |
|  | Missing | 7 | 7 |  |
|  | (95% CI proportion of difference) |  | (24.68%, 38.65%) |  |
| **Alleviation of all symptoms at 48h, n (%)** | Yes | 147 (42.2%) | 90 (25.4%) | <0.0001* |
|  | No | 201 (57.8%) | 265 (74.6%) |  |
|  | Missing | 4 | 0 |  |
|  | (95% CI proportion of difference) |  | (10.00, 23.77) |  |
| **Time to alleviation of systemic and respiratory flu-like symptoms (hours)** | Mean (SD) | 50.58 (24.55) | 69.23 (37.46) | <0.0001*** |
|  | Median (Q1-Q3) | 46.68 (35.0 – 64.9) | 67.17 (41.67 – 87.70) |  |
|  | KM estimate for the median time (95% CI) | 46.7 (44.00, 49.32) | 67.3 (60.08, 69.17) |  |
| **Time to return to usual daily activities (days)** | Mean (SD) | 4.5 (1.83) | 5.4 (2.26) | <0.0001*** |
|  | KM estimate for median time (95% CI) | 4.0 (NE) | 6.0 (5.00, 6.00) |  |
|  | Q1-Q3 | 3–6 | 3–7 |  |
| **Number of days of absence from work/school (days)** | Mean (SD) | 1.96 (1.52) | 2.23 (1.64) | 0.0218**** |
|  | Median (min, max) | 2 (0.0, 8.0) | 2 (0.0, 6.0) |  |
|  | Q1-Q3 | 1–3 | 1–3 |  |
|  | Missing | 54 | 34 |  |
| **Proportion of participants absent from work or school between the treatment groups, n (%)** | Yes | 237 (79.5%) | 264 (82.2%) | 0.3906* |
|  | No | 61 (20.5%) | 57 (17.8%) |  |
|  | Missing | 54 | 34 |  |

95% CI for KM estimate for median time to return to usual daily activities was not estimable (the standard error estimated based on Greenwood's formula was not stable).
\**P*-value was based on Chi-squared test.
\*\**P*-value was based on Cochran–Mantel–Haenszel test (to consider stratification).
\*\*\**P*-value based on stratified log-rank test.
\*\*\*\**P*-value was based on stratified Wilcoxon test.
95% CI for the proportion difference is based on normal approximation method without stratification.
CI = confidence interval; ILI = influenza-like illness; ITT = intention-to-treat; KM = Kaplan–Meier; min = minimum; max = maximum; N = number of participants in specified treatment group; n = number of participants in a specified category; NE = not estimable; Q1 = lower quartile; Q3 = upper quartile; SD = standard deviation.

#### 3.3.2. Time to alleviation of ILI symptoms

Median time to alleviation of systemic and respiratory flu-like symptoms was significantly shorter in the active treatment group compared with the placebo group (46.7 hours *vs.* 67.3 hours; *p* < 0.0001) **(Fig. 3**, **Table 4)**.

**Figure 3.**
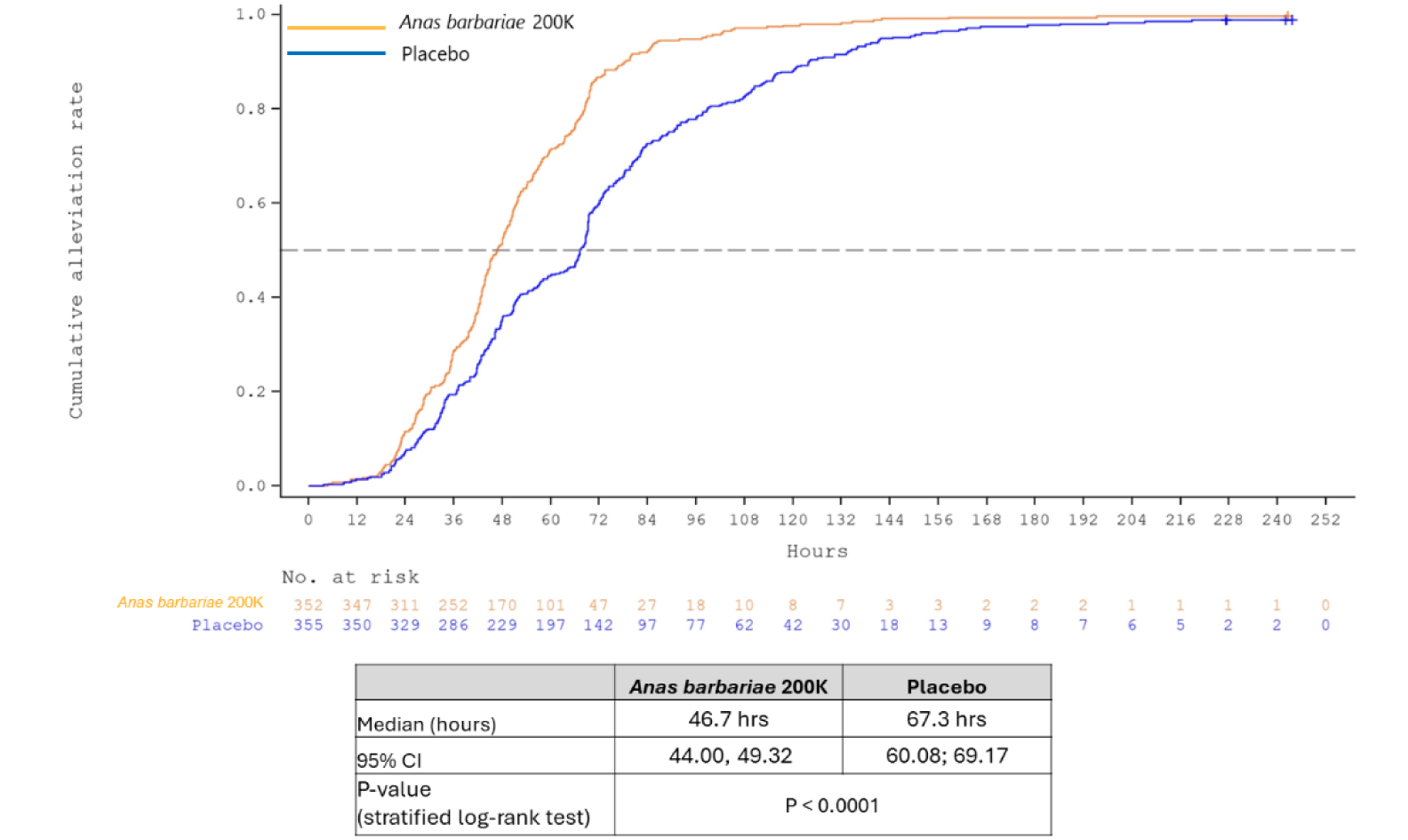
Kaplan–Meier plot for the cumulative distribution of time to alleviation of systemic and respiratory flu-like symptoms in the ITT analysis set. CI = confidence interval; ILI = influenza-like illness; ITT = intention-to-treat.

Specifically, median time to alleviation was significantly shorter for most individual symptoms; no significant differences were observed for fever and shortness of breath **(Table 5)**

**Table 5.** Summary of time to alleviation of systemic and respiratory flu-like symptoms and each symptom (ITT population)

| Statistic | *Anas barbariae* 200K<br>(N = 352) | Placebo<br>(N = 355) | *P*-value |
| --- | --- | --- | --- |
| **Time to alleviation of systemic flu-like symptoms (hours)** |  |  | <.0001 |
| Mean (SD) | 43.7 (24.27) | 57.1 (37.26) |  |
| KM estimate for median time (95% CI) | 41.3 (36.50, 43.62) | 47.8 (45.00, 51.70) |  |
| **Time to alleviation of respiratory flu-like symptoms (hours)** |  |  | <.0001 |
| Mean (SD) | 41.3 (23.33) | 54.7 (37.03) |  |
| KM estimate for median time (95% CI) | 40.7 (38.48, 42.68) | 46.2 (42.42, 51.20) |  |
| **Time to alleviation of fever (hours)** |  |  | 0.0532 |
| Mean (SD) | 18.7 (17.12) | 21.4 (20.19) |  |
| KM estimate for median time (95% CI) | 17.0 (13.50, 18.48) | 18.4 (15.93, 19.97) |  |
| **Time to alleviation of chills (hours)** |  |  | 0.0081 |
| Mean (SD) | 32.1 (24.22) | 39.4 (35.11) |  |
| KM estimate for median time (95% CI) | 25.1 (22.50, 29.67) | 25.5 (21.00, 32.05) |  |
| **Time to alleviation of malaise (hours)** |  |  | 0.0153 |
| Mean (SD) | 34.9 (27.48) | 41.1 (34.94) |  |
| KM estimate for median time (95% CI) | 25.8 (21.40, 33.50) | 32.6 (24.33, 41.08) |  |
| **Time to alleviation of headache (hours)** |  |  | 0.0104 |
| Mean (SD) | 35.7 (24.00) | 42.4 (33.03) |  |
| KM estimate for median time (95% CI) | 29.0 (26.83, 33.37) | 32.7 (27.00, 38.50) |  |
| **Time to alleviation of myalgia (hours)** |  |  | 0.0007 |
| Mean (SD) | 35.0 (25.28) | 45.4 (37.18) |  |
| KM estimate for median time (95% CI) | 29.0 (24.58, 34.45) | 36.6 (27.92, 43.50) |  |

| Statistic | *Anas barbariae* 200K<br>(N = 352) | Placebo<br>(N = 355) | P-value |
| --- | --- | --- | --- |
| **Time to alleviation of cough (hours)** |  |  | <.0001 |
| Mean (SD) | 35.5 (23.13) | 43.2 (31.77) |  |
| KM estimate for median time (95% CI) | 32.5 (27.67, 38.48) | 38.6 (33.83, 42.42) |  |
| **Time to alleviation of sore throat (hours)** |  |  | <.0001 |
| Mean (SD) | 34.9 (22.37) | 44.5 (34.83) |  |
| KM estimate for median time (95% CI) | 33.1 (27.42, 38.82) | 33.6 (27.75, 41.13) |  |
| **Time to alleviation of shortness of breath (hours)** |  |  | 0.1421 |
| Mean (SD) | 22.7 (29.07) | 26.1 (36.34) |  |
| KM estimate for median time (95% CI) | 18.4 (12.00, 21.98) | 18.4 (7.58, 23.03) |  |

Stratification factors were participant's age group and severity at baseline. Time was measured in hours.
P-value is based on stratified log-rank test.
CI = confidence interval; ITT = intention-to-treat; KM = Kaplan–Meier; N = number of participants in specified treatment group; SD = standard deviation.

#### 3.3.3. Absence from work/school

Median time to return to usual daily activities was significantly lower for the active treatment group compared with the placebo group (4.0 days *vs.* 6.0 days, *p* < 0.0001). The mean number of days of absence was significantly lower in the active treatment group compared with the placebo group (1.96 days *vs*. 2.23 days, *p* = 0.0218). However, there was no statistically significant difference in the proportion of participants who were absent from work or school between the treatment groups (79.5% *vs*. 82.2%, *p* = 0.3906) **(Table 4)**.

#### 3.3.4. Alleviation of other ILI symptoms

The active treatment produced a statistically significant greater reduction in fatigue compared to placebo, as evaluated at all scheduled timepoints from timepoint 5 through 23 (*p* < 0.05 for all comparisons). For the stuffy/runny nose symptom, statistically significant differences between the treatment groups were observed at timepoints 4 and 14, and all time points between 6 and 12 (*p* < 0.05 for each). At 72 hours (timepoint 9), the active treatment demonstrated a significant improvement compared to placebo in fatigue (83.5% *vs*. 74.6%, *p* = 0.0037) and stuffy/runny nose symptoms (92.3% *vs*. 85.9%, *p* = 0.0062). However, for gastrointestinal disorders, no significant differences were observed between the treatment groups, except at timepoint 5 **(Table 6).**

**Table 6.** Summary of alleviation of other ILI symptoms across multiple timepoints (ITT population)

| Symptom |  | *Anas barbariae* 200K<br>(N=352)<br>n (%) | Placebo<br>(N=355)<br>n (%) | P-value |
| --- | --- | --- | --- | --- |
| Fatigue | Timepoint 5 | 243 (69.0%) | 213 (60.0%) | 0.0103 |
|  | Timepoint 9 | 294 (83.5%) | 265 (74.6%) | 0.0037 |
|  | Timepoint 23 | 287 (81.5%) | 294 (82.8%) | 0.0489 |
| Nasal congestion | Timepoint 4 | 275 (78.1%) | 254 (71.5%) | 0.0440 |
|  | Timepoint 6 | 305 (86.6%) | 284 (80.0%) | 0.0178 |
|  | Timepoint 9 | 325 (92.3%) | 305 (85.9%) | 0.0062 |
|  | Timepoint 12 | 351 (99.7%) | 339 (95.5%) | 0.0002 |
|  | Timepoint 14 | 349 (99.1%) | 344 (96.9%) | 0.0321 |
|  | Timepoint 23 | 285 (81.0%) | 297 (83.7%) | 0.5408 |
| Gastrointestinal disorders | Timepoint 5 | 343 (97.4%) | 334 (94.1%) | 0.0219 |
|  | Timepoint 9 | 349 (99.1%) | 352 (99.2%) | 0.9917 |
|  | Timepoint 14 | 351 (99.7%) | 353 (99.4%) | 0.5679 |
|  | Timepoint 23 | 287 (81.5%) | 297 (83.7%) | 0.3260 |

ITT = intention-to-treat; N = number of participants in specified treatment group; n = number of participants in a specified category. P-value is based on Chi-squared/Fisher's Exact test.
Timepoint 4 (equivalent to 32 h); Timepoint 5 (equivalent to 40 h); Timepoint 6 (equivalent to 48 h) Timepoint 9 (equivalent to 72 h); Timepoint 12 (equivalent to 108 h; 4.5 days); Timepoint 14 (equivalent to 132 h; 5.5 days); Timepoint 23 (equivalent to 240 h; 10 days).

#### 3.3.5. Treatment satisfaction

Treatment satisfaction was assessed at the final evaluation using a study-specific 5-point ordinal scale (very poor, poor, neutral, good, or very good). Participants and investigators independently evaluated their satisfaction with the study treatment. High levels of treatment satisfaction were reported by the participants, with 88.9% (n = 313) rating the active treatment as “good” or “very good” compared with 35.8% (n = 127) for the placebo. Similar ratings were reported by investigators (88.6% (n = 312) *vs.* 40.6% (n = 144)) (**Supplemental Material, Table S.2**).

#### 3.3.6. Compliance and adverse events

High treatment adherence was observed, with 98.3% of participants in the active treatment group and 97.2% in the placebo group achieving 80%–120% compliance rates, as measured through participant diaries and reported doses (**Supplemental Material, Table S.3**).

A total of 25 participants (3.5%) experienced at least one AE during the study, with a comparable incidence between treatment groups (active treatment: n = 11 (3.1%); placebo: n = 14 (3.9%)). Only one moderate AE (diarrhea) was reported in the active treatment group and one mild AE (nasal congestion) in the placebo group was assessed as treatment-related. No SAE was reported **(Supplemental Material, Table S.4)**.

## 4. Discussion

This study, the largest to date evaluating *Anas barbariae* 200K, demonstrated significantly greater ILI symptom alleviation within 72 hours compared with placebo. Furthermore, participants receiving the active treatment experienced a significant 20-hour reduction in median time to symptom alleviation. Symptom alleviation varied according to baseline symptom severity. One possible explanation is that participants with moderate symptoms had the greatest opportunity to achieve a reduction of at least one severity category across all symptoms during follow-up, whereas participants with mild symptoms had limited room for improvement (ceiling effect) and those with severe symptoms required more substantial clinical improvement to meet the endpoint criteria. A similar proportion of absent participants across groups likely reflects enrolment timing, as participants may have already decided to be absent by the time of consultation. The active treatment effect was therefore observed in the duration of absence rather than its occurrence, consistent with the significantly shorter time to return to daily activities. Both participants and investigators reported greater treatment satisfaction with *Anas barbariae* 200K compared to placebo. Compliance was high and both treatments were well tolerated.

Consistent with the findings of Ferley *et al.* (1989) and Papp *et al.* (1998), all three placebo-controlled trials demonstrated statistically significant superiority of *Anas barbariae* 200K over placebo. Ferley *et al*. reported improved recovery within 48 hours (17.1% *vs.* 10.3%; *p* = 0.03)^24^, while Papp *et al*. observed a higher proportion of symptom-free participants at 48 hours (17.4% vs. 6.6%; *p* = 0.023)^25^. Direct comparison of response rates across studies should, however, be interpreted with caution because of important methodological differences. Previous studies evaluated outcomes at 48 hours, whereas symptom alleviation in the present trial was assessed at 72 hours. Endpoint definitions also differed, with Papp *et al*. using complete symptom resolution and this study defining symptom alleviation as an improvement of at least one severity category from baseline. Study populations and settings also differed, as the current trial enrolled pediatric participants in general practice settings across India, whereas earlier studies were conducted in European adult populations. These differences likely contributed to the higher response rates observed in both groups in the present study. The 72-hour assessment corresponded to the full treatment course, allowing evaluation of the complete therapeutic regimen.

Additional factors may include the broader ILI case definition used, cultural and healthcare-system differences, and the conduct of the study during the COVID-19 pandemic, which may have influenced healthcare-seeking behaviors and symptom reporting. Although differences in study design preclude direct quantitative comparisons, the findings remain directionally consistent across studies. Several systematic reviews and meta-analyses have evaluated homeopathy in acute conditions.^27–29^ Those studies reported findings suggestive of effects beyond placebo but highlighted methodological limitations and recommended the need for further high-quality trials to confirm the findings.

Despite the consistent clinical benefits observed, the mechanism of action of *Anas barbariae* 200K remains uncertain. Preliminary mechanistic evidence like the study by Paumier *et al.* (2025) showed that *Anas barbariae* 200K modulates microglial cell responses, reducing stiffness in inflamed cells and decreasing oxidative stress, including intracellular and mitochondrial reactive oxygen species. These findings provide preliminary insights into potential mechanisms associated with inflammation-related cellular processes.^30^

### Strengths and limitations

Randomization and blinding improved internal validity and reduced the risk of bias. The inclusion of a placebo control group enabled to discriminate the treatment specific effect from the placebo effect. The multicenter approach across general practice settings further strengthened the external validity and applicability of findings within the Indian primary care context. In addition, this study is the first randomized clinical trial evaluating *Anas Barbariae* 200K in this pediatric population, providing valuable data in an understudied population. The large sample size and extended follow-up timepoint allowed for a robust assessment of both short- and longer-term outcomes. The robustness of the results is further supported by their statistical significance in both the ITT and PP analyses. Moreover, the low dropout rate reflects strong participant adherence and follow-up quality. Potential confounding (vaccination) and mediating (paracetamol) factors were carefully considered and addressed in the analysis, allowing for a more accurate interpretation of the treatment effect and limiting residual confounding. Finally, the low number of AE observed reinforces the favorable safety profile of the treatment in the studied population.

Despite these strengths, several limitations warrant consideration. The absence of virological confirmation and reliance on self-reported outcomes may have reduced internal validity and introduced reporting bias. In addition, baseline symptom severity was classified using the participant’s most pronounced symptom and the investigator’s clinical judgment, without a validated severity grading instrument. This may have introduced variability in severity classification across investigators and could have influenced the interpretation of severity-dependent treatment effects. The study was conducted exclusively in general practice settings across India, limiting the generalizability of the findings to other healthcare systems, populations, or geographical regions. Adolescents aged 13–17 years were not included in the study population, limiting the applicability of the findings to this age group. Several factors may have contributed to the higher placebo response rate observed in this study relative to prior trials, including differences in study populations, the broader ILI case definition used, and outcome definitions. Influenza vaccination could not be included as a covariate due to the very low vaccination rate, leaving potential residual confounding unaddressed. Finally, the study was conducted during the COVID-19 pandemic, which may have influenced participant behavior and healthcare practices, potentially impacting the results of the study. Preventive measures and altered care-seeking patterns during this period could have affected symptom reporting and treatment outcomes, potentially limiting the generalizability of the findings.^31,32^

## 5. Conclusions

This large, randomized, placebo-controlled trial provides evidence supporting the efficacy of *Anas barbariae* 200K in alleviating ILI symptoms within 72 hours. *Anas barbariae* 200K showed statistically significant and clinically relevant improvements in both primary and most secondary efficacy outcomes. This homeopathic treatment may offer a valuable therapeutic option in the management of early ILI symptoms, with demonstrated safety, good tolerability, and strong participant and investigator-reported satisfaction. The clinical benefits demonstrated across several studies, combined with its established use in clinical practice, support consideration of *Anas barbariae* 200K as a therapeutic option for managing ILI symptoms.

## Supporting information

CONSORT checklist

## Data Availability

All data produced in the present study are available upon reasonable request to the corresponding author.

## Acknowledgements

We acknowledge the contributions of the participating general practitioners, patients and their families, as well as the clinical research staff involved in the study. The authors also recognize the statistical analysis support provided by Adrien Français, an employee of Delta Consultant. We appreciate Innovate Research team for project management and site relationship management. We thank IQVIA for project management and medical writing support, which was funded by Boiron, Messimy, France. We are grateful to Justine Vachon, PharmD. (Boiron) for project management and manuscript drafting and reviewing as well as Melissa Masmoudi, PhD. (Boiron), and Mathilde Savall, PhD. (Boiron) for their valuable assistance in reviewing this manuscript.

## Author contributions

**Manoj Gupta:** Conceptualization, Investigation, Formal analysis, Writing-review and editing and Visualization. **Ashish Tajne**: Investigation, Writing-review and editing and Visualization. **Pravin Chavan**: Investigation, Writing-review and editing and Visualization. **Isabelle Chanel**: Funding acquisition, Resources, Software, Supervision, Validation and Writing-review and editing. **Stéphanie Chanut**: Funding acquisition, Project administration, Resources, Software, Supervision, Validation, Writing-review and editing and Visualization. **Naoual Boujedaini**: Conceptualization, Funding acquisition, Methodology, Project administration, Resources, Supervision, Validation and Writing-review and editing. **Monica Gupta**: Conceptualization, Data curation, Formal analysis, Investigation, Writing-review and editing and Visualization.

All the authors are accountable for the work and provided their agreement for submission to the journal and approval for publication.

## Statements and declarations Ethical considerations

The protocol was reviewed and approved by the Royal Pune Independent Ethics Committee, the Independent Ethics Committee Healthpoint Hospital, the Institutional Ethics Committee Panchsheel Hospital, the Institutional Ethics Committee Narayana Diagnostic, and the Vagus Institutional Ethics Committee for this work. Ethics approval was granted in accordance with the Indian Council of Medical Research National Ethical Guidelines (2017, updated 2026). The study site and ethics committee details for participating investigators can be provided upon request from corresponding author.

## Consent to participate

All participants provided written informed consent prior to enrolment. The study was conducted in compliance with the Declaration of Helsinki, Good Clinical Practice guidelines, and applicable national regulations.

## Consent for publication

Not applicable.

