## Supplemental material for "Efficacy and safety of *Anas barbariae* 200K in influenza-like illness: a randomized double-blind, placebo-controlled, multicenter study"

1. Supplementary Methods

**Inclusion and exclusion criteria:**

Inclusion criteria:

- Aged 6 to 12 years for pediatrics' population or aged ≥ 18 years up to 65 years for adult population;
- Patients or parents of patients accepting to participate in the study through signing informed consent;
- Patients with ILI defined as sudden onset of symptoms and at least one of the following systemic symptoms: fever (≥37.8 °C/ 100.04 °F) or feverishness (feeling of fever or a chill) malaise headache, myalgia and at least one of the following respiratory symptoms: cough, sore throat, shortness of breath, of less than 24h hours duration;
- Patients able to take the first dose of study medication within 24 hours following the first symptoms of ILI.

Exclusion criteria:

- Patients and parents refusing to sign the informed consent;
- Patients with unstable and uncontrolled renal, cardiac, pulmonary, vascular, neurologic, metabolic (diabetes, thyroid disorders, adrenal disease), or immunodeficiency disorders, cancer, hepatitis, cirrhosis, asthma or chronic obstructive pulmonary disease (COPD);
- Participation in a clinical study with an investigational drug within 4 weeks prior to study entry;
- Patients who experienced a previous episode of acute upper respiratory tract infection (URTI), sinusitis, bronchitis, otitis or pneumonia within 4 weeks prior to the initial visit;
- Patients taking corticosteroids or immuno-suppressant therapies within 2 months prior to the initial visit;
- Patients with evidence/history of alcoholism, drug abuse, psychiatric disorders (including dementia or dementia like syndrome) or any other medical condition that could affect data collection;

- Treatment within 2 weeks prior to the initial visit with antiviral drugs such as neuraminidase inhibitors (Tamiflu®, Relenza®, Inavir®), or amantadine (Symmetrel®);
- Treatment within 1 week prior to initial visit with *Anas barbariae* 200K or antibiotics related to respiratory tract infection;
- Treatment within 1 week prior to initial visit with antipyretics other than paracetamol, analgesics, decongestants;
- Treatment within 1 week prior to initial visit with homeopathic medicine related to ILI at baseline or any other herbal medicine product or dietary supplement known to affect immune and/or inflammatory response;
- Patients with any other disease that requires immediate start of antibiotic treatment related to respiratory tract;
- Pregnant or breast-feeding women;
- Patients with intolerance of fructose, malabsorption of glucose or galactose, sucrase/isomaltase deficit;
- Any other condition which according to the investigator's judgement is not compatible with the principles of the study, e.g. inability to give informed consent, inability to complete the diary;
- Patients whom positive Covid-19 test.

**Statistical analysis**

- Sample size:

The sample size was calculated based on a one-sided alpha of 5% and 80% statistical power. Based on prior data[27], 80% of participants in the placebo group were expected to achieve symptoms alleviation at 72 hours. To detect a 7% absolute difference between groups (87% vs. 80%), which was considered clinically meaningful based on prior data, a total of 694 participants (347 per group) were required. Assuming a 10% dropout rate, the planned sample size was increased to 764 participants.

- Protocol deviations

Protocol deviation was pre-specified in the protocol and Statistical Analysis Plan. The review was conducted after database lock and unblinding according to predefined criteria.
Critical deviations included, for example, inclusion of a participant before obtaining a signed informed consent form. Major types of deviations included: 1) Violation of inclusion/exclusion criteria, 2) Non-compliance with study medication as failure to take medications at the prescribed times, and 3) Use of study-prohibited medications.

- Statistical analysis

All statistical analyses were conducted using SAS software version 9.4. The primary endpoint was analyzed using the chi-squared test and the Cochran–Mantel–Haenszel test with a multivariable regression model. Age, symptom severity, influenza vaccination status and paracetamol were initially considered as independent variables. However, influenza vaccination was excluded from the final model due to the very low vaccination rate. The final model adjusted for age, symptom severity, and paracetamol use.

Results included parameter estimates, *p*-values, odds ratios (OR), and two-sided 90% confidence intervals (CIs) for the primary analysis and one-sided 95% CIs for the secondary analysis. As this was a superiority test for the primary endpoint, one-sided testing at $\alpha = 0.05$ was used. Confidence intervals for the primary analysis are reported as two-sided 90% CIs, which are mathematically equivalent to the one-sided 95% CIs corresponding to the superiority hypothesis. One-sided 95% CIs were reported for the secondary analysis. Primary analyses were based on the observed-case dataset, with multiple imputation methods applied to handle missing data. The protocol prespecified one primary endpoint and several secondary endpoints. As no formal adjustment for multiplicity was planned, statistical inferences for secondary endpoints should be considered supportive and exploratory.

Time-to-event secondary outcomes were analyzed using stratified log-rank tests and Kaplan–Meier (KM) estimates. Categorical outcomes (e.g. absence from work or school) were compared using chi-square or Fisher's exact tests, while continuous variables (e.g., duration of absence) were analyzed using stratified Wilcoxon tests. A summary of treatment-related adverse events, including AEs categorized by preferred term and system organ class, as well as SAEs was reported for each treatment group.

2. Supplemental Tables

Table S.1 Usage of paracetamol (ITT population)

| **Statistic** | ***Anas barbariae* 200K (N = 352) n (%)** | **Placebo (N = 355) n (%)** | **ITT Population (N=707)** |
|---|---|---|---|
| Subjects used paracetamol | 193 (54.8%) | 198 (55.8%) | |
| Subjects did not used paracetamol | 159 (45.2%) | 157 (44.2%) | |
| **Paracetamol use (*Anas barbariae* 200K vs. Placebo)** | | | |
| *P*-value | | | 0.4002 |

ITT = intention-to-treat; N = number of participants in specified treatment group; n = number of participants in a specified category. *P*-value is based on Chi-square test.

Table S.2 Summary of participant and investigator satisfaction (ITT population)

| **Category** | ***Anas barbariae* 200K (N = 352) n (%)** | **Placebo (N = 355) n (%)** |
|---|---|---|
| **Investigator satisfaction** | | |
| Very Good or Good | 312 (88.6%) | 144 (40.6%) |
| Very Good | 114 (32.4%) | 29 (8.2%) |
| Good | 198 (56.3%) | 115 (32.4%) |
| Neutral | 37 (10.5%) | 138 (38.9%) |
| Poor | 1 (0.3%) | 62 (17.5%) |
| Very Poor | 2 (0.6%) | 11 (3.1%) |
| **Participant satisfaction** | | |
| Very Good or Good | 313 (88.9%) | 127 (35.8%) |
| Very Good | 119 (33.8%) | 10 (2.8%) |
| Good | 194 (55.1%) | 117 (33.0%) |
| Neutral | 35 (9.9%) | 160 (45.1%) |
| Poor | 3 (0.9%) | 63 (17.7%) |
| Very Poor | 1 (0.3%) | 5 (1.4%) |

ITT = intention-to-treat; N = number of participants in specified treatment group; n = number of participants in a specified category.

Table S.3 Summary of compliance with study drug (ITT population)

| **Compliance level** | ***Anas barbariae* 200K (N = 352) n (%)** | **Placebo (N = 355) n (%)** |
|---|---|---|
| <80% | 6 (1.7%) | 10 (2.8%) |
| 80%–120% | 345 (98.3%) | 345 (97.2%) |
| >120% | 0 | 0 |
| Missing | 1 | 0 |

ITT = intention-to-treat; N = number of participants in specified treatment group; n = number of participants in a specified category.

Table S.4 Summary of adverse events (Safety population)

| **Category** | ***Anas barbariae* 200K (N = 352) n (%)** | **Placebo (N = 355) n (%)** | **Overall (N = 707) n (%)** |
|---|---|---|---|
| With at least one AE | 11 (3.1%) | 14 (3.9%) | 25 (3.5%) |
| Respiratory, thoracic and mediastinal disorders | 3 (0.9%) | 9 (2.5%) | 12 (1.7%) |
| Gastrointestinal disorders | 5 (1.4%) | 3 (0.8%) | 8 (1.1%) |
| Metabolism and nutrition disorders | 2 (0.6%) | 2 (0.6%) | 4 (0.6%) |
| Uncoded | 1 (0.3%) | 0 | 1 (0.1%) |
| With at least one SAE | 0 | 0 | 0 |
| With at least one TRAE | 0 | 1 (0.3%) | 1 (0.1%) |
| With at least one treatment-related severe AE | 0 | 0 | 0 |
| With at least one treatment-related SAE | 0 | 0 | 0 |
| With at least one AE leading to discontinuation of study drug | 0 | 0 | 0 |
| With at least one AE leading to death due to study drug | 0 | 0 | 0 |

AE = adverse event; N = number of participants in specified treatment group; n = number of participants in a specified category; SAE = serious adverse event; TRAE = treatment-related adverse event.

The number of events was identical to the number of subjects reporting the event for all categories; therefore, only n (%) are presented.
